# Serum Slit3 is associated with disease activity and interstitial lung disease in rheumatoid arthritis

**DOI:** 10.1101/2025.01.05.25320017

**Authors:** Xuepei Zhang, Maohua Shi, Dongmei Guo, Yangtao Yu, Hongwei Zhang, Guoqiang Chen

**Affiliations:** 1Department of Rheumatology, First People’s Hospital of Foshan, Foshan 528000, China; 2Department of Rheumatology, Second People’s Hospital of Foshan, Foshan 528000, China

**Keywords:** Rheumatoid arthritis, Slit guidance ligand 3, Disease activity, Inflammation, Interstitial lung disease

## Abstract

Rheumatoid arthritis (RA) is a chronic systemic inflammatory disease accompanied by high incidence rate of interstitial lung disease (ILD). Slit guidance ligand 3 (Slit3) which is a member of axon guidance molecule superfamily, mediates a series of biological activities. In this study, we explored the potential clinical significance of Slit3, especially as a serum biomarker for evaluating RA-ILD. Patients diagnosed with RA were recruited between July 2022 to April 2024 from the Department of Rheumatology of Foshan First People’s Hospital. The clinical data were collected from hospital records, serum level of Slit3 was detected by enzyme-linked immunosorbent assay (ELISA). Among 232 recruited patients, RA patients in high Slit3 subgroup were older, showed higher proportion of positive rheumatoid factor (RF) status and anti-cyclic citrullinated peptide antibody (APCA) status, as well as higher levels of ESR, CRP, Disease Activity Score in 28 joints with four variables, including C-reactive protein (DAS28-CRP) or erythrocyte sedimentation rate (DAS28-ESR), Simplified Disease Activity Index (SDAI) and pro-inflammatory cytokine interleukin-6 (IL-6). Notably, Logistic regression analysis showed that elevated serum Slit3 was an independent relevant factor of RA-ILD (OR 1.005 [95% CI 1.002-1.008], P=0.002). Our findings evealed that serum Slit3 was associated with disease activity in RA and may hold promise as a useful serum biomarker and therapeutic target for RA-ILD.

## Introduction

Rheumatoid arthritis (RA) is a chronic systemic inflammatory disease characterized by progressive and erosive destruction of arthritis, accompanied by damage to multiple organs^[1]^. Interstitial lung disease (ILD), the most common extra-articular complication of RA, clinically affects up to 10% of patients with RA and subclinically affects up to 40%. What’s more, RA-ILD is the driving cause of death in patients with RA, leading to significant morbidity and mortality^[2,3]^. Identifying factors that are associated with RA-ILD would help precisely manage and improve prognosis of these patients.

Slit guidance ligand 3 (Slit3) which is a member of axon guidance molecule superfamily, is a highly conserved secreted glycoprotein widely expressed in various tissues. By binding to their receptor Robos, Slits activate intracellular signaling pathways in order to mediate a series of biological activities inculding axonal rejection, neuronal migration, organ development, reproductive regulation, tumor metastasis and angiogenesis, et al^[4,5]^. It was reported that Slit3 enhanced monocyte migration in a concentration-dependent and chemoattractant-independent manner, contributed myeloid leukocyte recruitment to the inflamed peritoneum in vivo^[6]^. Slit3 is highly expressed in cardiac fibroblasts and elevated migration and differentiation of rat cardiac fibroblasts, as well as fibrillar collagen production, making it a potential therapeutic target for fibrotic diseases^[7,8]^. These data showed that Slit3 may be a key regulator of inflammatory response and ILD. However, the expression and clinical significance of Slit3 in RA have not been reported.

In this study, we first analyzed the expression of Slit3 in the peripheral blood of RA patients, and continued to explore the clinical significance of Slit3, especially as a serum biomarker for evaluating RA-ILD.

## Methods

### Data Sources

RA patients aged ≥ 16 years and fulfilled the 2010 American College of Rheumatology (ACR) / European League Against Rheumatism (EULAR) classification criteria for RA^[9]^ were recruited between July 2022 to April 2024 from the Department of Rheumatology of Foshan First People’s Hospital, China. Exclusion criteria were as follows: overlapping other autoimmune diseases (e.g. systemic lupus erythematosus, scleroderma, and dermatomyositis), serious infection, malignancy and pregnancy.

Healthy control subjects were enrolled from the Physical Examination Center of the hospital. The Ethics Committee of Foshan First People’s Hospital approved this study, and all subjects signed informed consent forms before clinical data collection.

### Clinical assessments

The demographic and clinical data were collected from hospital records that could identify individual participants during or after data collection. The data included age, gender, disease duration, disease activity, physical function, radiographic indicators and medications.

Disease duration was divided into three categories: <6 months (short), 6-24 months (intermediate), and >24 months (long). Physical function using the Health Assessment Questionnaire Disability Index (HAQ-DI) indicators, as we described previously ^[10]^.

Disease activity was assessed using the Disease Activity Score in 28 joints with four variables, including C-reactive protein (DAS28-CRP) or erythrocyte sedimentation rate (DAS28-ESR), the Simplified Disease Activity Index (SDAI), and the Clinical Disease Activity Index (CDAI). Disease activity, as defined by DAS28-CRP, was divided into four categories: high disease activity (DAS28-CRP > 5.1), moderate disease activity (3.2 ≤ DAS28-CRP ≤ 5.1), low disease activity (2.6 ≤ DAS28-CRP < 3.2), and remission (DAS28-CRP < 2.6) ^[11,12]^.

Conventional radiographs of bilateral hands and wrists (anteroposterior view) were assessed with the Sharp/van der Heijde modified score^[13]^ as we described previously^[10]^. ILD was diagnosed based on high-resolution computed tomography (HRCT) findings, demanding fibrotic abnormality in over 10% of lung parenchyma, optionally with traction bronchiectasis or honeycombing, and with no evidence or suspicion of an alternate diagnosis^[14]^.

### Serum Slit3 measurement

Serum samples of RA patients and healthy controls were collected after overnight fasting and stored at –80°C. The serum level of Slit3 was measured using a commercially available enzyme-linked immunosorbent assay (ELISA) kit (NeoBioscience Technology Co.,Ltd., OKDD00532) according to the manufacturer’s instructions.

### Statistical analysis

Statistical analyses were performed with SPSS software (version 25.0). Data were summarized by means ± standard deviations (SD) or medians with interquartile range (IQR) for continuous variables according to the data distributions and the frequencies with percentages for categorical variables. The Mann-Whitney test was used to compare the differences of continuous variables between two groups. The Chi-square test or Fisher exact test was used for categorical variables in two groups. Univariate and multivariate logistic regression analyses were used to identify potential associated factors of RA-ILD. All significance tests were two-tailed and were conducted at the 5% significance level.

## Results

### Baseline characteristics of the study patients

There were totally 232 RA patients and 50 healthy control subjects recruited (Table 1). In RA group, the median age was 62.0 (54.3,71.0) years with 73.7% female. The median disease duration was 48 (12,120) months, 15.9% with short disease duration (<6 months) and 59.5% with long disease duration (>24 months) . According to DAS28-CRP, there were 27.6%, 43.1%, 11.6% and 17.7% patients in high, moderate, low disease activity and remission, respectively. There were 20.3% patients without previous glucocorticoid or conventional synthetic disease-modifying anti-rheumatic drugs (DMARDs) therapy for at least six months before enrolment (treatment naive).

**Table 1.**
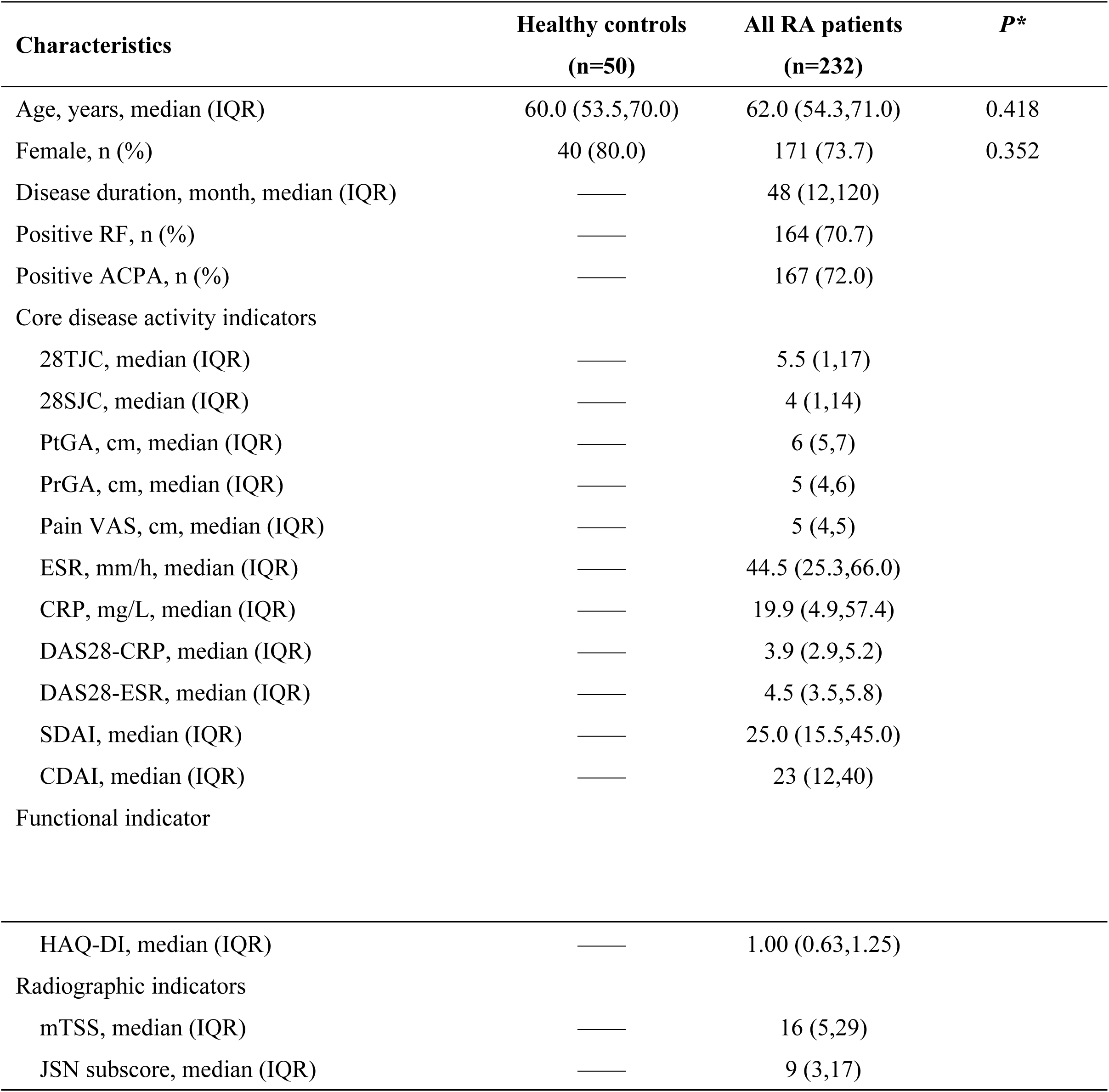
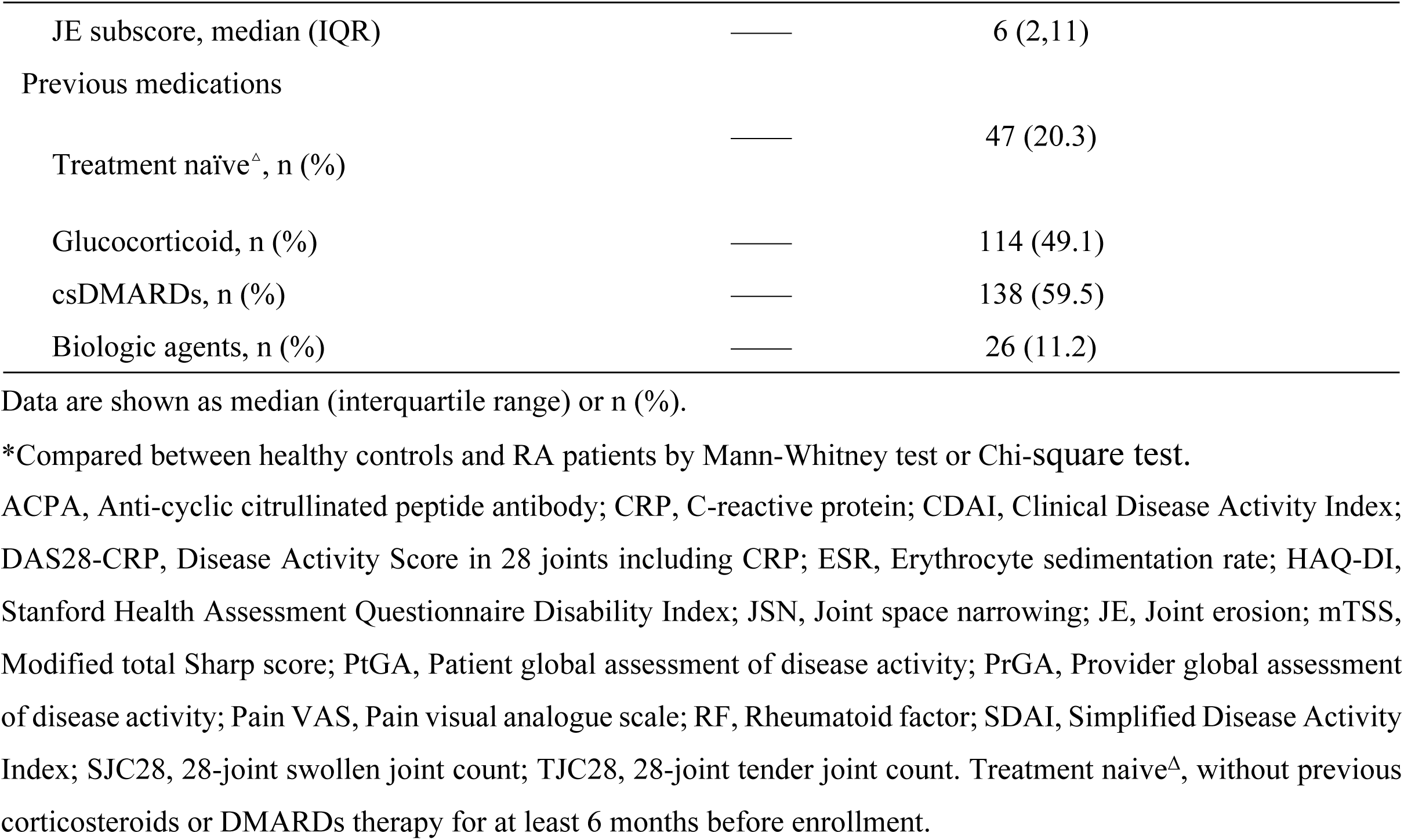
Baseline characteristics of RA patients and healthy controls.

| Characteristics | Healthy controls<br>(n=50) | All RA patients<br>(n=232) | *P* * |
| --- | --- | --- | --- |
| Age, years, median (IQR) | 60.0 (53.5,70.0) | 62.0 (54.3,71.0) | 0.418 |
| Female, n (%) | 40 (80.0) | 171 (73.7) | 0.352 |
| Disease duration, month, median (IQR) | — | 48 (12,120) |  |
| Positive RF, n (%) | — | 164 (70.7) |  |
| Positive ACPA, n (%) | — | 167 (72.0) |  |
| Core disease activity indicators |  |  |  |
| 28TJC, median (IQR) | — | 5.5 (1,17) |  |
| 28SJC, median (IQR) | — | 4 (1,14) |  |
| PtGA, cm, median (IQR) | — | 6 (5,7) |  |
| PrGA, cm, median (IQR) | — | 5 (4,6) |  |
| Pain VAS, cm, median (IQR) | — | 5 (4,5) |  |
| ESR, mm/h, median (IQR) | — | 44.5 (25.3,66.0) |  |
| CRP, mg/L, median (IQR) | — | 19.9 (4.9,57.4) |  |
| DAS28-CRP, median (IQR) | — | 3.9 (2.9,5.2) |  |
| DAS28-ESR, median (IQR) | — | 4.5 (3.5,5.8) |  |
| SDAI, median (IQR) | — | 25.0 (15.5,45.0) |  |
| CDAI, median (IQR) | — | 23 (12,40) |  |
| Functional indicator |  |  |  |
| **Table 1. (Continued)** |  |  |  |
| HAQ-DI, median (IQR) | — | 1.00 (0.63,1.25) |  |
| Radiographic indicators |  |  |  |
| mTSS, median (IQR) | — | 16 (5,29) |  |
| JSN subscore, median (IQR) | — | 9 (3,17) |  |

|  |  |  |
| --- | --- | --- |
| JE subscore, median (IQR) | — | 6 (2,11) |
| Previous medications |  |  |
| Treatment naïve<sup>Δ</sup>, n (%) | — | 47 (20.3) |
| Glucocorticoid, n (%) | — | 114 (49.1) |
| csDMARDs, n (%) | — | 138 (59.5) |
| Biologic agents, n (%) | — | 26 (11.2) |

Data are shown as median (interquartile range) or n (%).
\*Compared between healthy controls and RA patients by Mann-Whitney test or Chi-square test.
ACPA, Anti-cyclic citrullinated peptide antibody; CRP, C-reactive protein; CDAI, Clinical Disease Activity Index; DAS28-CRP, Disease Activity Score in 28 joints including CRP; ESR, Erythrocyte sedimentation rate; HAQ-DI, Stanford Health Assessment Questionnaire Disability Index; JSN, Joint space narrowing; JE, Joint erosion; mTSS, Modified total Sharp score; PtGA, Patient global assessment of disease activity; PrGA, Provider global assessment of disease activity; Pain VAS, Pain visual analogue scale; RF, Rheumatoid factor; SDAI, Simplified Disease Activity Index; SJC28, 28-joint swollen joint count; TJC28, 28-joint tender joint count. Treatment naïve<sup>Δ</sup>, without previous corticosteroids or DMARDs therapy for at least 6 months before enrollment.

In healthy control group (n=50), the median age was 60.0 (53.5,70.0) with 80% female. There was no difference in age or gender between healthy control subjects and RA patients (both *P*>0.05 ).

### Serum Slit3 level in RA patients

Compared with healthy controls, RA patients showed a higher level of serum Slit3 [182.0 (78.7-296.6) ng/ml *vs.* 89.4 (61.4-143.0) ng/ml, *P*<0.001, Fig 1A]. Further gender stratification analysis showed that both female and male RA patients had higher levels of serum Slit3 [female: 183.7 (79.4-284.6) ng/ml *vs.* 92.1 (66.1-144.1) ng/ml, *P*<0.001, Fig 1B; male: 181.7 (67.3-317.0) ng/ml *vs.* 79.0 (55.5-149.2) ng/ml, *P*=0.040, Fig 1C].

**Fig 1.**
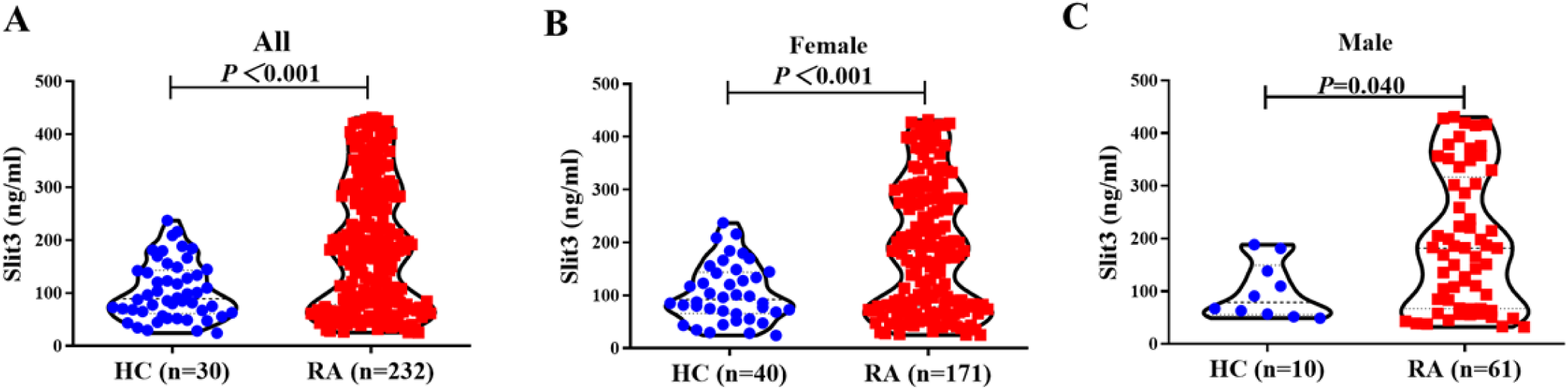
Serum Slit3 level in RA patients and healthy controls. Baseline serum Slit3 level in RA patients and healthy controls. HC, healthy controls; RA, rheumatoid arthritis.

### Correlation between serum Slit3 with disease characteristics

According to the median level of serum Slit3 (182.0ng/ml), RA patients were stratified into two subgroups including low Slit3 group (n=116) and high Slit3 group (n=116). Compared the characteristics between the two subgroups, RA patients in high Slit3 group were older (median 65 *vs.* 59), showed higher proportion of positive rheumatoid factor (RF) status (76.7% *vs.*64.7% ) and anti-cyclic citrullinated peptide antibody (APCA) status (80.2% *vs.*63.8%), as well as higher level of disease activity indicators including ESR (median 56 *vs.* 36), CRP (median 29.6 *vs.* 10.6), DAS28-CRP (median 4.2 *vs.* 3.6), DAS28-ESR (median 4.8 *vs.* 4.2) and SDAI (median 31.5 *vs.* 23.1, all *P*<0.05, Table 2).

**Table 2.** Characteristics of RA patients in serum Slit3 subgroups.

|  | Low Slit3 group<br>(n=116) | High Slit3 group<br>(n=116) | *P* |
| --- | --- | --- | --- |
| Age, years, median (IQR) | 59 (53,67) | 65 (56,73) | **0.008** |
| Female, n (%) | 87 (75.0) | 84 (72.4) | 0.655 |
| Disease duration, mo, median (IQR) | 48 (12,120) | 48 (12,120) | 0.864 |
| Positive RF, n (%) | 75 (64.7) | 89 (76.7) | **0.043** |
| Positive ACPA, n (%) | 74 (63.8) | 93 (80.2) | **0.005** |
| Core disease activity indicators |  |  |  |
| 28TJC, median (IQR) | 4 (1,16) | 8 (2,18) | 0.056 |
| 28SJC, median (IQR) | 2 (0,16) | 5 (1,14) | 0.214 |
| PtGA, median (IQR) | 5 (4,6) | 6 (5,7) | 0.057 |
| PrGA, median (IQR) | 5 (4,6) | 5 (4,6) | 0.017 |
| Pain VAS, median (IQR) | 5 (3,5) | 5 (4,6) | 0.058 |
| ESR (mm/h), median (IQR) | 36.0 (19.0,57.0) | 56.0 (33.0,72.0) | **<0.001** |
| CRP (mg/L), median (IQR) | 10.6 (2.9,40.9) | 29.6 (7.5,71.0) | **<0.001** |
| DAS28-CRP, median (IQR) | 3.6 (2.6,5.0) | 4.2 (3.5,5.4) | **0.003** |
| DAS28-ESR, median (IQR) | 4.2 (3.2,5.4) | 4.8 (3.9,5.9) | **0.003** |
| SDAI, median (IQR) | 23.1 (12.9,41.0) | 31.5 (19.0,46.7) | **0.016** |
| CDAI, median (IQR) | 18 (11,38) | 29 (14,41) | 0.068 |
| Functional assessment |  |  |  |
| HAQ-DI, median (IQR) | 0.8 (0.5,1.1) | 1.1 (0.6,1.3) | **0.001** |
| Radiographic assessments |  |  |  |
| mTSS, median (IQR) | 15 (5,31) | 16 (5,28) | 0.995 |
| JSN subscore, median (IQR) | 11 (2,17) | 9 (3,16) | 0.776 |
| **Table 2. (Continued)** |  |  |  |
| JE subscore, median (IQR) | 5 (2,12) | 6 (2,11) | 0.623 |
| Previous medications |  |  |  |
| Treatment-naïve<sup>A</sup>, n (%) | 27 (23.3) | 20 (17.2) | 0.253 |
| Corticosteroids, n (%) | 51 (44.0) | 63 (54.3) | 0.115 |
| csDMARDs, n (%) | 65 (56.0) | 73 (62.9) | 0.285 |
| Biologic agents, n (%) | 15 (12.9) | 11 (9.5) | 0.405 |

Data are shown as median (interquartile range) or n (%).
\*Compared between RA patients in low slit3 subgroup and high slit3 subgroup by Mann-Whitney test or Chi-square test.
Significant differences are shown in bold.
ACPA, Anti-cyclic citrullinated peptide antibody; CRP, C-reactive protein; CDAI, Clinical Disease Activity Index; DAS28-CRP, Disease Activity Score in 28 joints including CRP; ESR, Erythrocyte sedimentation rate; HAQ-DI, Stanford Health Assessment Questionnaire Disability Index; JSN, Joint space narrowing; JE, Joint erosion; mTSS, Modified total Sharp score; PtGA, Patient global assessment of disease activity; PrGA, Provider global assessment of disease activity; Pain VAS, Pain visual analogue scale; RF, Rheumatoid factor; SDAI, Simplified Disease Activity Index; SJC28, 28-joint swollen joint count; TJC28, 28-joint tender joint count. Treatment naive<sup>Δ</sup>, without previous corticosteroids or DMARDs therapy for at least 6 months before enrollment.

Furthermore, the correlations between serum Slit3 level with different serum cytokine levels examined in 75 RA patients were analyzed. Results showed a significant positive correlation between serum Slit3 and pro-inflammatory cytokine interleukin-6 (IL-6) (r=0.336, *P*=0.003, Fig 2), but not with other cytokines including IL-4, IL-10, IL-12p70, IL-17,tumor necrosis factor (TNF) and r-transcriptional intermediate factor (rTIF) .

**Fig 2.**
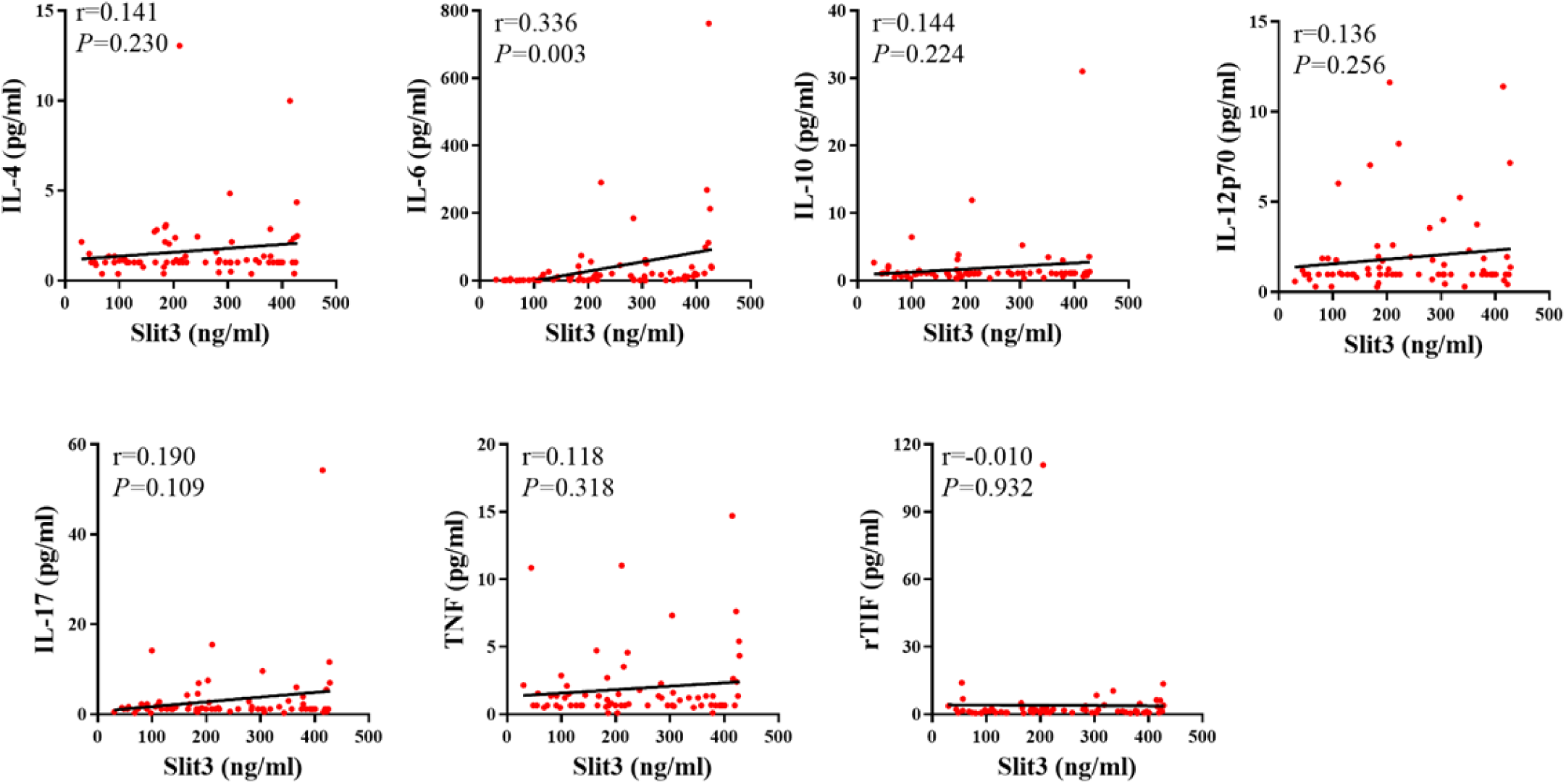
Correlation between serum Slit3 with different serum cytokine levels.

### The relevant characteristics of RA-ILD

Among the 232 RA patients, there were 45 (19.4%) RA patients showing ILD. Compared with those without ILD (RA-no-ILD), RA patients with ILD were older (median 66 *vs.* 62), showed lower proportion of female (51.1% *vs.*79.1%), higher proportion of positive RF status (88.9% *vs.*66.3%) and APCA status (91.1% *vs.*67.4%), higher levels of ESR (median 59 *vs.* 42) and HAQ-DI (median 1.1 *vs.* 0.9), as well as higher level of serum Slit3 (median 259.5 *vs.* 168.8, all *P*<0.05, Table S1).

Univariate logistic regression analysis showed that RA-ILD was positively associated with high serum Slit3 [odds ratio (OR)=1.005, 95% CI 1.002-1.007], male gender [OR=3.630, 95% CI 1.834-7.184], long disease duration [OR=1.003, 95% CI 1.000-1.006], positive RF status [OR=4.065, 95% CI 1.529-10.807], positive ACPA status [OR=4.962, 95% CI 1.700-14.485] and high ESR [OR=1.015, 95% CI 1.004-1.027, all *P*<0.05]. Further stepwise multivariate logistic regression analysis, including the significant indicators mentioned above, showed that high serum Slit3 [odds ratio (OR)=1.005, 95% CI 1.002-1.008], male gender [OR=5.561, 95% CI 2.498-12.380], long disease duration [OR=1.005, 95% CI 1.002-1.009], positive ACPA status [OR=3.608, 95% CI 1.161-11.218, all *P*<0.05 ] were the independent characteristics associated with RA-ILD (Fig 3).

**Fig 3.**
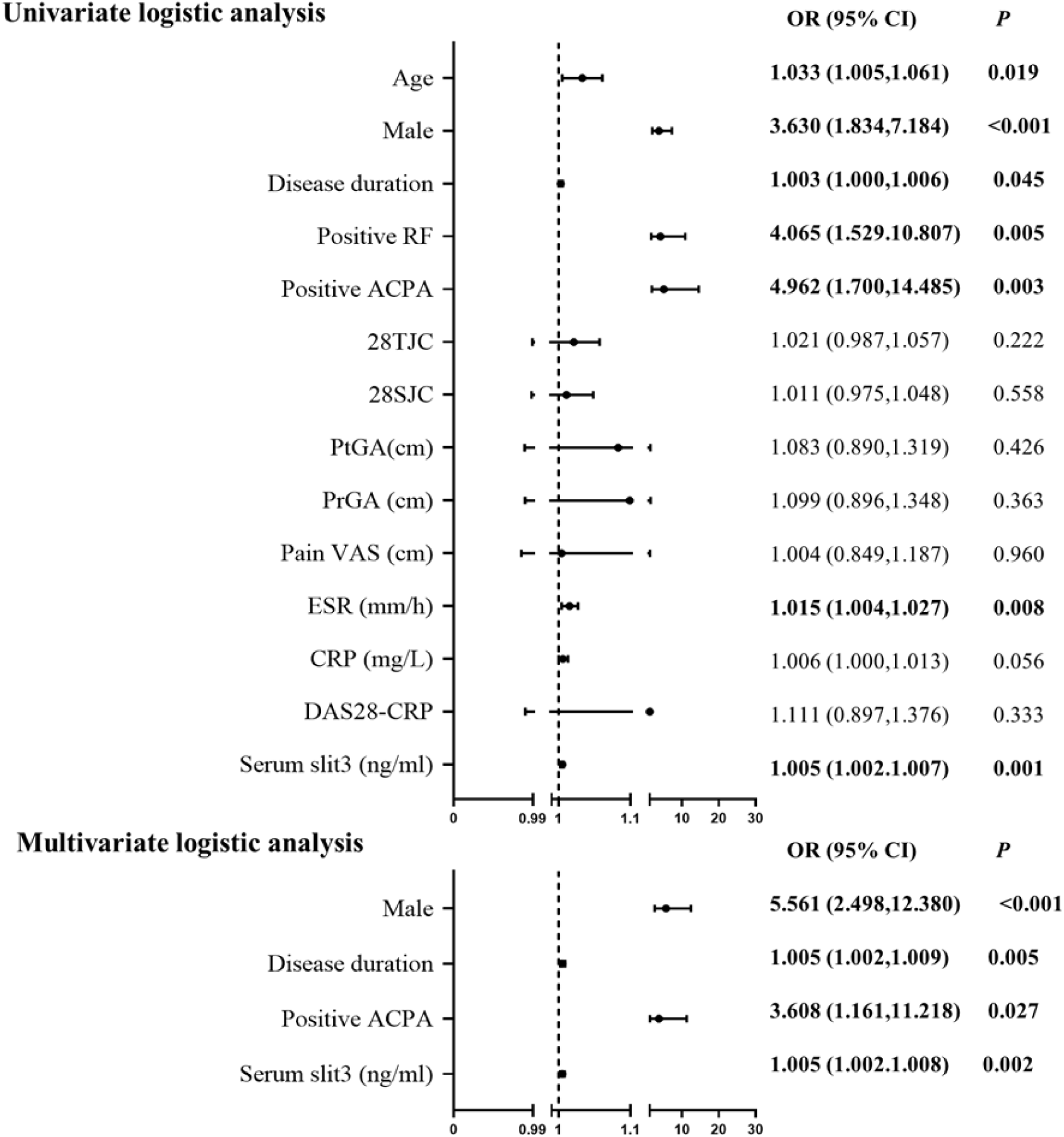
Logistic regression analysis of the relevant characteristics of RA-ILD. ACPA, Anti-cyclic citrullinated peptide antibody; CRP, C-reactive protein; DAS28-CRP, Disease Activity Score in 28 joints including CRP; ESR, Erythrocyte sedimentation rate; PtGA, Patient global assessment of disease activity; PrGA, Provider global assessment of disease activity; Pain VAS, Pain visual analogue scale; RF, Rheumatoid factor; SJC28, 28-joint swollen joint count; TJC28, 28-joint tender joint count; OR, odds ratio; CI, confidence interval.

## Discussion

In this study, we investigated the relationship of serum Slit3 and disease characteristics in RA patients. RA patients with high serum Slit3 showed higher disease activity, higher proportion of ILD, and Slit3 is an independent risk factor of RA-ILD. All these findings indicted that serum Slit3 may be a critical regulator of inflammation and ILD in RA.

Slit3 is a highly conserved secreted glycoprotein expressing in many organs and is present at high level in human fibroblasts, tenocytes, adipocytes, mesenchymal cells and smooth muscle cells^[4,15]^. In the majority of tissue microenvironments, Slit3 can bind to Robo receptors to transmit cellular signals and regulate various life activities^[4,5]^. Previous studies showed that Slit3 acts as an angiogenic factor, stimulating endothelial cell proliferation, motility, chemotaxis, and formation of endothelial vascular network through interacting with Robo4^[16]^. Silencing of Slit3 promoted proliferation, migration, and invasion of lung adenocarcinoma, and treatment with Slit3 lead to inhibiting migration of malignant melanoma cells^[17,18]^.Recent researches showed that osteoclast-derived Slit3 plays an osteoprotective role by synchronously stimulating bone formation and suppressing bone resorption through combinating Robo receptors, making it a potential therapeutic target for metabolic bone disorders^[19,20]^. Another study showed that Slit2 and Slit3 mRNA levels were higher than Slit1 mRNA levels in RA synovial fibroblasts (RA-SF), comparing RA-SF and normal SF, RA-SF showed lower expression of Slit3, and Slit3 inhibits Robo3-induced migratory activity of RA-SF^[21]^. As for our study, compared with healthy controls, RA patients showed a higher level of serum Slit3 in both women and men. More researches are needed to explore the the main source of Slit3 in RA.

Previous studies have identified the link between Slit3 and inflammation. Stimulation with Slit3 increased the spontaneous and chemoattractant-induced migration of primary monocytes in vitro and increased the myeloid cell recruitment during peritoneal inflammation in vivo^[6]^. The total amounts and concentrations of Slit3 were significantly higher in periodontitis than those in healthy, and significant positive correlations were observed between the level of Slit3 in the gingival crevicular fluid and radiographic bone loss^[22]^. Elevated levels of Slit3 expression in amniotic and myometrium cells in preterm birth lead to rupture of the fetal membrane by promoting gene expression and release of IL-1β-induced pro-inflammatory cytokines (IL-6 and IL-8) and matrix metalloproteinase-9 (MMP9)^[23]^. Calculating high- and low-inflammatory indices of 490 lung adenocarcinoma patients in Cancer Genome Atlas (TCGA) database, the gene of Slit3 in the high inflammatory index group was upregulated, and patients with high expression of Slit3 showed satisfactory prognosis^[24]^. In our study, RA patients in high serum slit3 group showed higher disease activity indicators including ESR、CRP、DAS28-CRP、DAS28-ESR and SDAI. Slit3 also showed significant correlation with pro-inflammatory cytokine IL-6. All these indicated the pro-inflammatory effect of Slit3 in RA.

RA-ILD is the driving cause of death in patients with RA, leading to significant morbidity and mortality. While it can be the initial presenting manifestation in 10% to 20% of patients, most RA-ILD occurs within the first five years after diagnosis^[25]^.The assessment of risk factors is crucial for RA-ILD due to the disease’s relevant prevalence as well as the effect that the diagnosis has on mortality rates and treatment decisions. Studies showed that some significant human leukocyte antigen (HLA) variations including HLA-DRB1, HLA-DR4, and HLA-B40 can contribute to the emergence of ILD in RA patients. Antibodies including RF and ACPA, lung epithelial-related proteins such as Lungen-6 (KL-6),Surfactant protein D (SP-D), as well as cytokines such as CCL18, IL-6 and others such as MMP-7, ESR, CRP are also important biomarkers for RA-ILD^[26,27,28]^. A study showed that compared with RA who do not have ILD, those have ILD were more frequently men, older and had higher DAS28-ESR^[29]^. Similarly, in our study, male gender, long disease duration and positive ACPA status were the independent risk factors of RA-ILD.

Apart from inflammatory cascades, fibroblast proliferation and activation is also the main driver of ILD^[30]^. According to an analysis of GEO data, Slit3 is up-regulated in the liver tissue of people with fibrosing non-alcoholic steatohepatitis, and Slit3 expression level was positively correlated with fibrosis-related proteins. Human umbilical cord-derived mesenchymal stem cells are capable of alleviating liver fibrosis by inhibiting the activation of hepatic stellate cell (HSCs) through the miR-148a-5p/SLIT3 pathway^[31,32]^. Another study showed that Slit3 expression level was increased in Ang II-induced mice models and cardiac fibroblasts and promote cardiac fibrosis and fibroblast differentiation via the RhoA/ROCK1 signaling pathway^[33]^. These data imply the importance of Slit3 as a potential therapeutic target in tissue fibrosis^[34]^. However, another study showed that in mice, injections of Slit2 inhibit bleomycin-induced lung fibrosis. In lung tissue from pulmonary fibrosis patients with relatively normal lung function, Slit2 has a widespread distribution whereas, in patients with advanced disease, there is less Slit2 in the fibrotic lesions^[35]^. In our study, high serum Slit3 was the independent risk factor of RA-ILD, the mechanism of Slit3 in RA-ILD and lung fibrosis is worth further discussion.

The present study also had some limitations. Firstly, it was designed as a cross-sectional investigation. Risk factors and outcome measurement at the same timeframe made it scientifically inappropriate to determine the causality. A future large scale of multi-community based epidemiological survey on general population and multi-center prospective studies on RA patients are needed to address these limitations in our study and make potentially novel discoveries. Secondly, as the small sample, we didn’t analyze the factors associated with different subtypes of ILD, more samples need to be included to further classify the subtypes of ILD and explore the relationship of serum Slit3 as well as other risk factors with RA-ILD subtypes in the future study.

## Conclusion

In conclusion, in this study, we reported that elevated serum Slit3 was associated with higher disease activity and inflammation in RA, especially as an independent risk factor for RA-ILD. Slit3 may hold promise as a useful serum biomarker and therapeutic target for RA-ILD.

## Data Availability

All relevant data are within the manuscript and its Supporting Information files.

## Supporting information

**S1 Table. Comparison of characteristics between RA-ILD and RA-no-ILD. (PDF)**

## Author Contributions

**Conceptualization:** Guo-qiang Chen.

**Data curation:** Guo-qiang Chen.

**Formal analysis:** Xue-pei Zhang, Mao-hua Shi, Dong-mei Guo.

**Investigation:** Xue-pei Zhang, Mao-hua Shi, Dong-mei Guo, Yang-tao Yu.

**Validation:** Guo-qiang Chen, Xue-pei Zhang, Hong-wei Zhang.

**Writing-original draft:** Xue-pei Zhang.

**Writing-review & editing:** Guo-qiang Chen, Xue-pei Zhang, Mao-hua Shi, Dong-mei Guo, Yang-tao Yu, Hong-wei Zhang.

